# Changes in opioid prescribing during the COVID-19 pandemic in England: cohort study of 20 million patients in OpenSAFELY-TPP

**DOI:** 10.1101/2024.02.23.24303238

**Authors:** Andrea L Schaffer, Colm D Andrews, Andrew D Brown, Richard Croker, William J Hulme, Linda Nab, Jane Quinlan, Victoria Speed, Christopher Wood, Milan Wiedemann, Jon Massey, Peter Inglesby, Seb CJ Bacon, Amir Mehrkar, Chris Bates, Ben Goldacre, The OpenSAFELY Collaborative, Alex J Walker, Brian MacKenna

## Abstract

**Background:** The COVID-19 pandemic disrupted healthcare delivery, including difficulty accessing in-person care, which may have increased the need for strong pharmacological pain relief.

**Methods:** With NHS England approval, we used routine clinical data from >20 million general practice adult patients in OpenSAFELY-TPP. Using interrupted time series analysis, we quantified prevalent and new opioid prescribing prior to the COVID-19 pandemic (January 2018-February 2020), and during lockdown (March 2020-March 2021) and recovery periods (April 2021-June 2022), overall and stratified by demographics (age, sex, deprivation, ethnicity, geographic region) and to people in care homes.

**Outcomes:** The median number of people prescribed an opioid per month was 50.9 per 1000 patients prior to the pandemic. There was little change in prevalent prescribing during the pandemic, except for a temporary increase in March 2020. We observed a 9.8% (95%CI -14.5%, -6.5%) reduction in new opioid prescribing from March 2020, sustained to June 2022 for all demographic groups except people 80+ years. Among care home residents, in April 2020 new opioid prescribing increased by 112.5% (95%CI 92.2%, 134.9%) and parenteral opioid prescribing increased by 186.3% (95%CI 153.1%, 223.9%).

**Interpretation:** New opioid prescribing increased among older people and care home residents, likely reflecting use to treat end-of-life COVID-19 symptoms, but decreased among most other groups. Further research is needed to understand what is driving the reduction in new opioid prescribing and its relation to changes to health care provision during the pandemic.

**Funding:** The OpenSAFELY Platform is supported by grants from the Wellcome Trust (222097/Z/20/Z) and MRC (MR/V015737/1, MC_PC_20059, MR/W016729/1). In addition, development of OpenSAFELY has been funded by the Longitudinal Health and Wellbeing strand of the National Core Studies programme (MC_PC_20030: MC_PC_20059), the NIHR funded CONVALESCENCE programme (COV-LT-0009), NIHR (NIHR135559, COV-LT2-0073), and the Data and Connectivity National Core Study funded by UK Research and Innovation (MC_PC_20058) and Health Data Research UK (HDRUK2021.000). The views expressed are those of the authors and not necessarily those of the NIHR, NHS England, UK Health Security Agency (UKHSA) or the Department of Health and Social Care.

**Evidence before this study:** We searched Pubmed for publications between 1 March 2020 and 8 January 2023 using the following search terms: (“COVID-19” OR “SARS-CoV-2”) AND (“United Kingdom” OR “England” OR “Britain” OR “Scotland” OR “Wales”) AND (“opioid”). We also searched the reference list of relevant articles. We included research studies (excluding conference abstracts and editorials) that quantified opioid prescribing or use in the United Kingdom during the COVID-19 pandemic. Studies focussed solely on opioid substitution therapy for treatment of opioid use disorder were excluded.

We identified four studies. One described opioid use among a cohort of people on a waiting list for hip or knee arthroplasty in Scotland (n=548) and found higher rates of long-term opioid use during the COVID-19 pandemic compared with historical controls. The second study quantified changes in opioid prescribing using English aggregate prescription data. This study found no changes in opioid prescribing after the start of the COVID-19 pandemic. The third study of 1.3 million people with rheumatic and musculoskeletal diseases found a decrease in new opioid users among people with certain conditions, but not in the number of overall prescriptions. The last study of 34,711 people newly diagnosed with cancer and 30,256 who died of cancer in Wales found increases in strong opioid prescribing in both populations.

**Added value of this study:** This is the largest study (>20 million patients) of opioid prescribing during the COVID-19 pandemic in a representative sample of the population of England. We used person-level data to quantify changes in the number of people prescribed opioids and identified that prevalent opioid prescribing changed little, with the exception of a temporary increase at the start of the first lockdown. However, we also identified meaningful reductions in new opioid prescribing. While our findings confirm previous studies quantifying variation in opioid prescribing by sex, ethnicity, region and deprivation, we showed that changes to new prescribing during the COVID-19 pandemic were experienced approximately similarly across these subgroups. The exceptions were older people and people in care homes. The latter group experienced substantial increases in new opioid prescribing (especially parenteral opioids, which are used in palliative care) coinciding with periods of greatest COVID-19 morbidity and mortality.

**Implications of all the available evidence:** The COVID-19 pandemic resulted in substantial disruptions to the healthcare system. Despite concerns that difficulty or delays in providing care during the pandemic may have led to shifts from non-pharmacological treatments to greater opioid prescribing, we observed no increases in prescribing prevalence in most demographic groups in England. The one major exception is people residing in care homes, where the observed prescribing patterns suggest use to treat end of life symptoms, consistent with best practice. However, our findings do not preclude increased prescribing in high risk subgroups, such as people on procedure waiting lists. Further research to quantify changes in this population is warranted.

## Background

In England, 13% of adults received an opioid prescription in 2017/18.(1) While opioids are effective at treating acute pain, cancer pain, and end-of-life pain, they are commonly overprescribed for chronic non-cancer pain,(2) where opioids lack evidence of efficacy(3,4) and are not recommended.(5) During the COVID-19 pandemic, there were disruptions to provision of healthcare, including access to medicines, primary care appointments, and elective procedures. These disruptions were not experienced equally, with women, ethnic minorities, and older people most impacted,(6) the same populations disproportionately affected by opioid-related harms.(7,8)

International studies quantifying opioid prescribing during the COVID-19 pandemic have identified changes not consistent with best practice. A Canadian study found increases in prescribing to people living in care homes,(9) a population known to be at high risk of opioid-related harms. A US study identified a shift from non-pharmacological treatment (e.g. physical therapy) towards opioid therapy for people with pain,(10) likely due to increasing remote care during the pandemic. Furthermore, data suggest that rates of opioid-related death and overdose were greater than expected during the COVID-19 pandemic in Canada.(11)

While changes in prescribing have been observed during COVID-19 in the UK for different classes of medicines(12), or in specific populations(13,14), there are no studies on changes to opioid prescribing at the person-level in the general population or in high-risk demographic groups. Due to the risks associated with overprescribing of opioids, especially to vulnerable populations, we set out to quantify changes to the following measures during the COVID-19 pandemic, overall and by key subgroups: 1) prevalent opioid prescribing; 2) new prescribing; 3) variation in COVID-19-related changes by demographic subgroups and people in care homes.

## Methods

### Study design

We conducted an interrupted time series analysis study from January 2018 to June 2022. We defined two change points, the start of the “restrictions period”, defined as March 2020 as the UK first introduced restrictions on 26 March, and the start of the “recovery period”, defined as April 2021. April 2021 was chosen as it coincides with the start of gradual reopening of non-essential services.(15)

### Data source and data sharing

Primary care records managed by the GP software provider, TPP were linked to Office of National Statistics (ONS) death data through OpenSAFELY and were linked, stored and analysed securely within the OpenSAFELY platform: https://opensafely.org/ as part of the NHS England OpenSAFELY COVID-19 service. Data include pseudonymised data such as coded diagnoses, medications and physiological parameters. No free text data are included. All code is shared openly for review and re-use under MIT open license (https://github.com/opensafely/opioids-covid-research). Detailed pseudonymised patient data are potentially re-identifiable and therefore not shared.

### Study population

We identified all people prescribed an opioid in each month of the study period (January 2018 to June 2022). All people aged 18 years or older, alive, and registered with a TPP practice on the first of every month were included in the denominator for calculation of rates. We excluded people with missing data on age and sex.

### Study measures

Opioids were defined as all medicines falling under the British National Formulary (BNF) legacy paragraphs 4.7.2 (Opioid analgesics), as well as opioid medicines falling under 3.9.1 (Cough suppressants), and opioid-containing combination medicines under 4.7.1 (Non-opioid analgesics),1.4.2 (Antimotility drugs), and 10.1.1 (Non-steroidal antiinflammatory drugs). Opioids used to treat opioid use disorder were not included. Links to the codelists used in this study are openly available for inspection and re-use in this study’s Github repository (https://github.com/opensafely/opioids-covid-research).

### Population characteristics

We characterised all people prescribed an opioid between the last three months of the study period (April-June 2022). Opioid prescribing rates were expressed as the number of people prescribed an opioid per 1000 registered adult patients. To prevent disclosure, all counts have been rounded to the nearest 7.

We included the following demographic categories: sex (male, female); age (18-39, 40-49, 50-59, 60-69, 70-79, 80-89, 90+ years); Index of Multiple Deprivation (IMD) deciles; practice region (East, East Midlands, London, North East, North West, South East, South West, West Midlands, Yorkshire and the Humber); and ethnicity (White [British, Irish, Other]; Asian or Asian British [Bangladeshi, Indian, Pakistani, Other]; Black or Black British [African, Caribbean, Other]; Mixed [White/Asian, White/Black African, White/Black Caribbean, Other]; Other [Chinese, Other]). To compare overall prescribing rates within relevant demographic categories (e.g., ethnicity, IMD decile, region) we standardised opioid prescribing rates by age (5-year age bands) and sex using the ONS mid-year 2020 English population.(16)

We identified people residing in care homes (a vulnerable population during the pandemic) based on a combination of coded events (e.g., identification of consultations that occurred in care homes) and by linking individuals’ registered address to those of care homes as held by the Care Quality Commission, further refined by applying the algorithm described by Schultze et al.(17)

### Opioid prescribing during the COVID-19 pandemic

#### Prevalent and new opioid prescribing

Opioid prescribing prevalence was defined as the number of people prescribed an opioid, and included both new and repeat prescriptions. We estimated changes in monthly prevalent opioid prescribing during the restriction and recovery periods using interrupted time series analysis. We used the crude (unstandardised) rates for these analyses, as relative changes which would not be affected by standardisation, and due to the additional lack of precision of standardised estimates.

We modelled the number of people prescribed an opioid using negative binomial log-linear regression, and included the natural log of the number of registered patients in each month as an offset. The model included variables representing the pre-COVID-19 trend (slope), as well as a level shift (i.e. an immediate, sustained change in prescribing) and a slope change (i.e. a gradual change in trend) after the start of both periods (restriction and recovery). We calculated Newey-West standard errors to account for residual autocorrelation and included dummy month variables to account for seasonality. Due to previous reports of increases in opioid sales in March 2020 followed by decreases in April and May 2020(18), we tested the base model described above for inclusion of dummy variables representing these individual months, which were retained if they improved model fit as determined by the likelihood ratio test. For each model we estimated incidence rate ratios (IRRs) and 95% confidence intervals (CIs) which were expressed as percent changes.

New opioid prescribing was defined as people prescribed an opioid without any opioid prescription in the previous year. We quantified changes as described for overall prescribing. However, here the offset was the number of opioid-naive registered patients in each month (i.e., people without any opioid prescription in the previous year).

As most concerns over opioid prescribing focus on use for chronic non-cancer pain, we repeated the above analyses excluding people with a cancer diagnosis in the past 5 years as a sensitivity analysis.

#### Opioid prescribing by formulation

We identified prescribing of high-dose long-acting opioids which are not recommended for chronic non-cancer pain.(3) Among long-acting opioids, high dose opioids were defined as those with ≥120 mg morphine equivalents per day based on the typical total daily dose, a definition used previously.(8) We also identified prescribing of parenteral opioids (i.e. delivered by injection or intravenously), recommended to treat end of life symptoms such as pain or breathlessness in the community.(19) The latter were included as we hypothesised that an an increase in COVID-19 mortality would be associated with an increase in medicines used in palliative care. We constructed interrupted times series models for each opioid formulation separately.

#### People living in care homes

We quantified changes in overall prescribing, prescribing of high-dose long-acting opioids, and parenteral opioids, among people living in care homes using the same approach as described above. For the outcome of high-dose long-acting prescribing to people in care homes, we used Poisson log-linear regression (instead of negative binomial) as this was a better fitting model.

#### Demographic subgroups

To estimate differences in overall and new opioid prescribing by demographic subgroups, we created separate models for each variable (age, sex, IMD decile, ethnicity, and region). People with missing values were excluded from this analysis (except for ethnicity). We used the base models established above and tested an interaction term between the level shift and change in slope variables and the categories of each variable. As inclusion of an interaction with the change in slope did not improve model fit for any of the subgroups, this interaction term was not retained. We therefore assumed a common trend and that the change in slope did not vary across groups, and only the level shift varied. Using these models, we estimated the change (level shift) in prescribing during the restriction and recovery periods for each subgroup.

### Software and reproducibility

Data management was performed using Python 3.8, with analysis carried out using R 4.0.5. Code for data management and analysis as well as codelists archived online.

### Patient and public involvement

We have involved patients and the public in various ways: we developed a public website that provides a detailed description of the platform in language suitable for a lay audience (https://opensafely.org); we have participated in two citizen juries exploring public trust in OpenSAFELY; we co-developed an explainer video; we have patient representation who are experts by experience on our OpenSAFELY Oversight Board; we have partnered with Understanding Patient Data to produce lay explainers on the importance of large datasets for research; we have presented at various online public engagement events to key communities; and more. To ensure the patient voice is represented, we are working closely to decide on language choices with appropriate medical research charities.

### Ethics approval

This study was approved by the Health Research Authority (Research Ethics Committee reference 20/LO/0651) and by the London School of Hygiene and Tropical Medicine Ethics Board (reference 21863).

## Results

### Population characteristics

From April to June 2022, there were 20,476,680 registered patients (≥18 years) with 1,445,122 prescribed an opioid, or 70.6 per 1000 registered patients. Opioid prescribing increased with age, ranging from 12.6 per 1000 people aged 18-29 years to 202.8 per 1000 people aged 90+ years (**Table 1**). Prescribing also increased with greater deprivation varying more than two-fold, ranging from 47.7 per 1000 for people in the least deprived IMD decile to 102.6 per 1000 for the most deprived. However, after age and sex standardisation these differences widened further, ranging from 42.0 per 1000 to 120.2 per 1000.

**Table 1.**
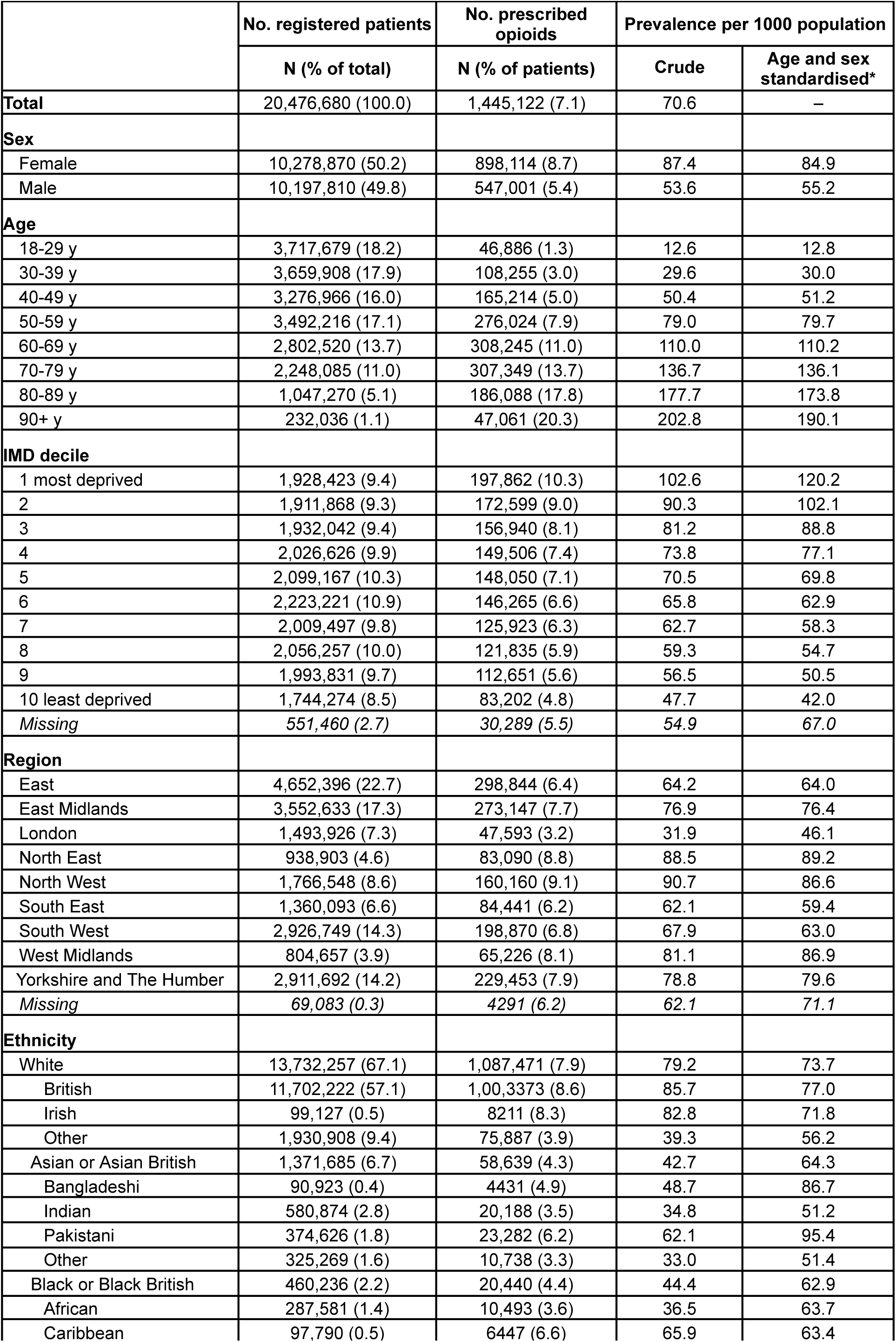
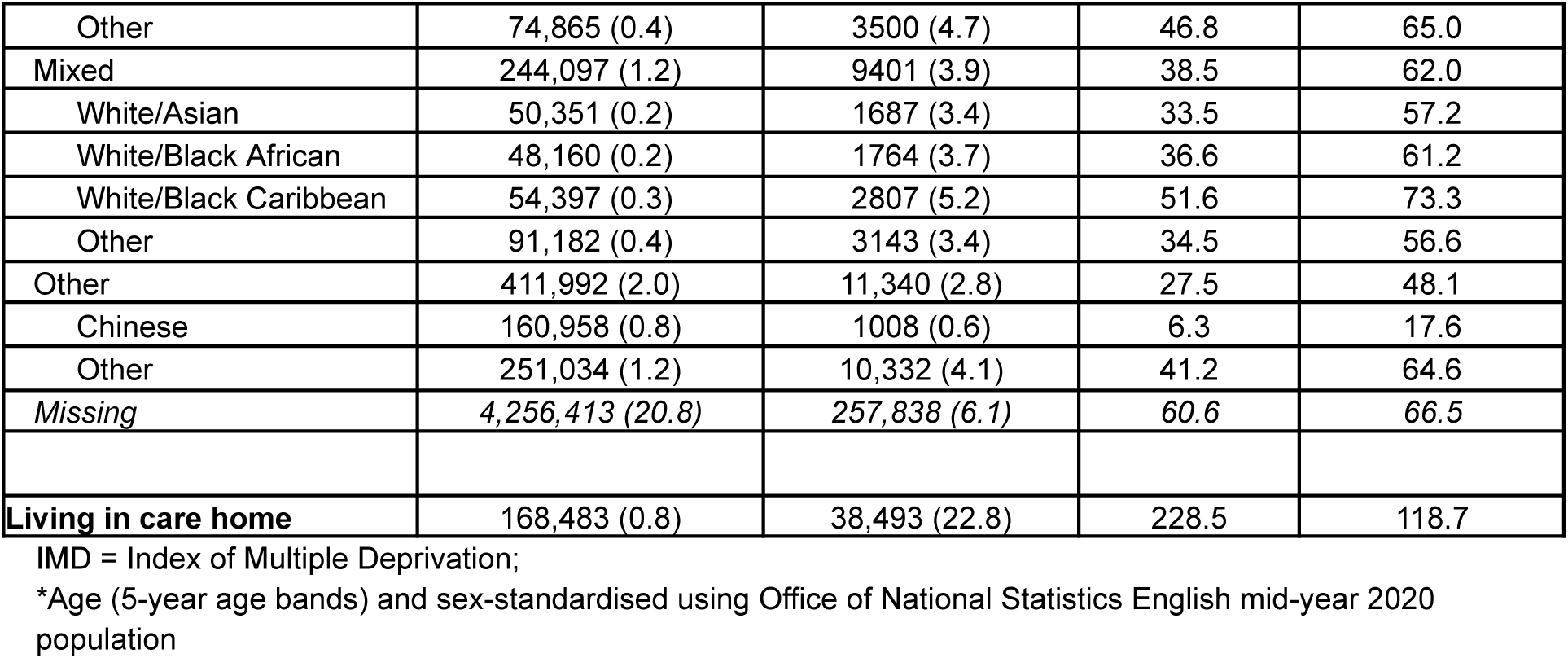
Registered adult patients (≥18 years) prescribed an opioid between April and June 2022. All counts rounded to nearest 7.

Age and sex standardised rates of opioid prescribing were also high in women (84.9 per 1000), people with Pakistani, Bangladeshi, White British and White Irish ethnicity (95.4, 86.7, 77.0 and 71.8 per 1000), and people living in the North East, North West and West Midlands (89.2, 86.6 and 86.9 per 1000). Differences by ethnicity were attenuated after age and sex standardisation. Among people residing in care homes (0.8% of all registered adult patients), nearly 1 in 4 were prescribed an opioid during this period (228.5 per 1000).

### Changes in opioid prescribing

#### Overall prescribing prevalence

There were 19,113,668 registered adult patients in January 2018 increasing over the study period to 20,510,959 in June 2022 (**Supplementary Figure 1**). The median prevalence of opioid prescribing was 50.9 per 1000 adult patients per month prior to COVID-19, and was declining by an estimated 0.3% per month (95%CI -0.3%, -0.2%) (**Figure 1**, **Table 2**). In March 2020, opioid prescribing prevalence was 7.0% higher than predicted had previous trends continued (95%CI 3.3%, 10.9%); this was followed by lower than expected rates in May (-4.7%, 95%CI -7.7%, -1.6%). Aside from these temporary pulses, no changes to the level or slope were observed during the restriction or recovery periods. Similar results were observed when excluding people with a cancer diagnosis in the past 5 years (**Supplementary Figure 2, Supplementary Table 1**).

**Figure 1.**
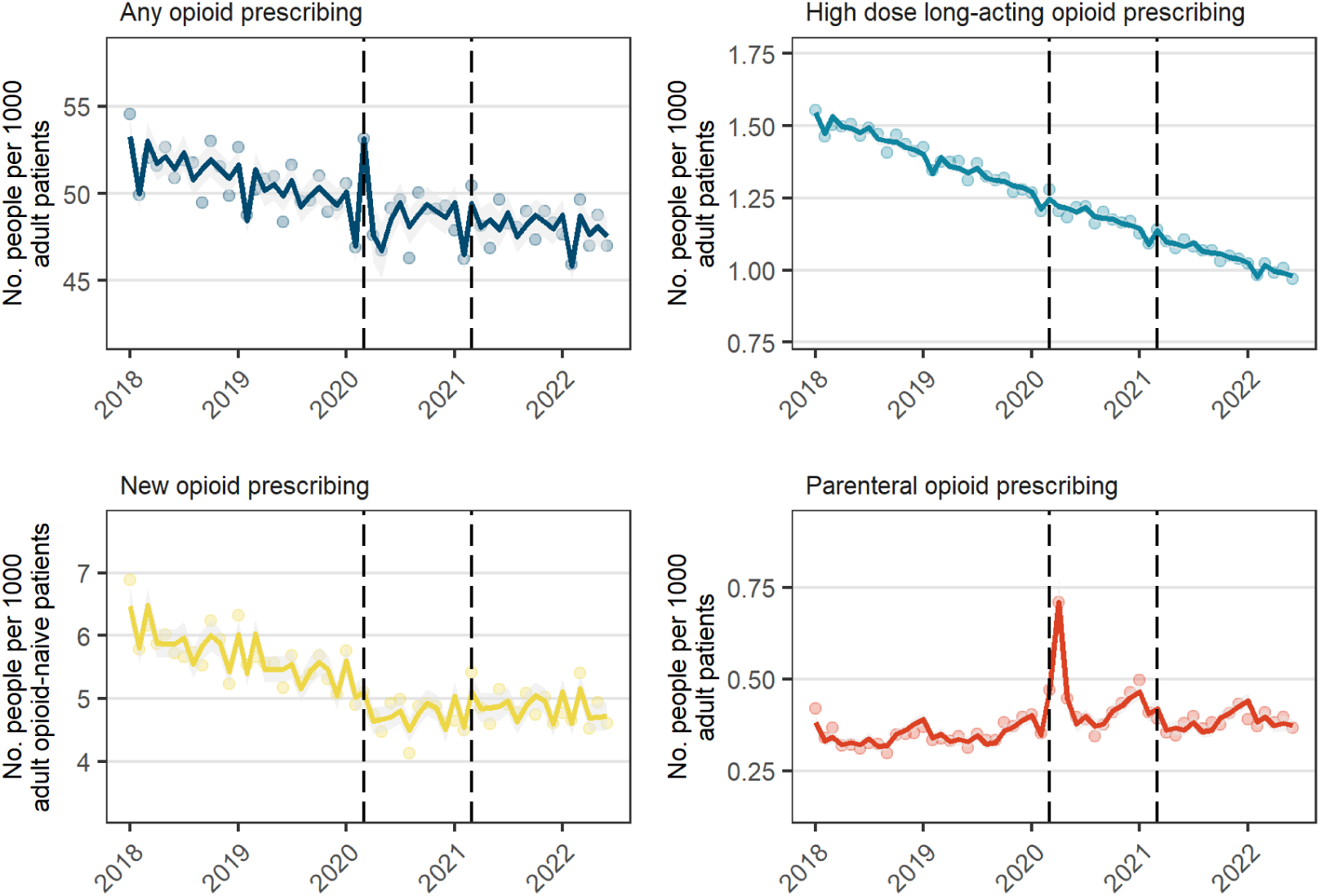
Number of people prescribed opioids per month (Jan 2018 to June 2022) among all registered adult patients. Solid lines are fitted values, dots are observed values, and vertical dashed lines represent start of restriction period (Mar 2020) and recovery period (Apr 2021).

**Table 2.** Relative changes in number of people prescribed opioids per 1000 population during the restriction (Mar 2020-Mar 2021) and recovery (Apr 2021-Jun 2022) periods among all registered adult patients and people living in care homes.

|  | Pre-COVID-19<br>monthly slope<br>(%, 95% CI) | Changes during restriction period relative to pre-COVID-19 |  |  |  |  | Changes during recovery period<br>relative to restriction period |  |
| --- | --- | --- | --- | --- | --- | --- | --- | --- |
|  |  | Level shift<br>(%, 95% CI) | Change in<br>slope (%, 95%<br>CI) | March 2020 (%,<br>95% CI) | April 2020 (%,<br>95% CI) | May 2020 (%,<br>95% CI) | Level shift (%,<br>95% CI) | Change in slope<br>(%, 95% CI) |
| **Full adult population** |  |  |  |  |  |  |  |  |
| Any opioid | -0.3 (-0.3, -0.2) | -0.6 (-3.4, 2.5) | 0.2 (-0.1, 0.6) | 7.0 (3.3, 10.9) | -2.0 (-4.2, 0.3) | -4.7 (-7.7, -1.6) | -0.6 (-3.0, 1.9) | -0.03 (-0.4, 0.3) |
| New opioid | -0.6 (-0.7, -0.5) | -9.8 (-14.5, -6.5) | 0.6 (0.2, 1.1) | * | * | * | 4.1 (-0.9, 9.4) | -0.3 (-0.8, 0.2) |
| High dose long-acting<br>opioid | -0.8 (-0.9, -0.8) | -1.1 (-2.6, 0.5) | 0.03 (-0.1, 0.2) | * | * | * | -1.2 (-2.5, 0.1) | -0.03 (-0.2, 0.1) |
| Parenteral opioid | 0.2 (-0.1, 0.5) | 10.7 (-0.4, 23.1) | 0.2 (-0.8, 1.3) | 18.0 (6.1, 31.2) | 89.4 (76.0, 103.8) | 16.8 (8.3, 26.0) | -8.4 (-14.6, -1.8) | -0.2 (-1.5, 1.1) |
| **People living in care**<br>**homes** |  |  |  |  |  |  |  |  |
| Any opioid | -0.2 (-0.3, -0.2) | 0.2 (-2.1, 2.4) | 0.09 (-0.2, 0.3) | 2.9 (0.3, 5.6) | 13.3 (11.2, 15.4) | 0.3 (-2.3, 2.9) | -1.5 (-3.0, -0.01) | 0.05 (-0.2, 0.3) |
| New opioid | -0.3 (-0.6, 0.04) | 4.4 (-9.4, 20.4) | 0.5 (-1.0, 2.1) | 12.9 (-2.3, 30.6) | 112.5 (92.2,<br>134.9) | 26.0 (14.6,<br>38.5) | -10.2 (-18.7, -0.7) | -0.06 (-1.8, 1.7) |
| High dose long-acting<br>opioid | -1.3 (-1.4, -1.1) | 1.7 (-1.0, 4.5) | -0.4 (-0.7, -0.2) | * | * | * | -0.6 (-2.9, 1.7) | 1.5 (1.3, 1.8) |
| Parenteral opioid | 0.03 (-0.5, 0.5) | -4.1 (-21.1, 16.6) | 1.2 (-1.1, 3.6) | 35.9 (11.6, 65.5) | 186.3 (153.1,<br>223.9) | 54.2 (37.0,<br>73.7) | -14.3 (-26.7, 0.2) | -0.8 (-3.5, 2.0) |

\*Not included in model

#### New opioid prescribing

There was a median of 5.7 people newly prescribed opioids per 1000 opioid-naïve patients per month and was declining by 0.6% per month pre-COVID-19 (95%CI -0.7%, -0.5%) (**Figure 1**, **Table 2**). In contrast to prevalent prescribing, no increase was observed in March 2020. Starting during the restriction period, there was a -9.8% level shift in new prescribing (95%CI -14.5%, -6.5%) and a 0.6% increase in slope (95%CI 0.2%, 1.1%) and weak evidence of a small upward shift during the recovery period (4.1%, 95%CI -0.9%, 9.4%).

#### By opioid formulation

High-dose long-acting opioids represented a small minority of opioid prescribing. The median prescribing prevalence was 1.4 per 1000 per month pre-COVID-19 and was declining by 0.8% per month (95%CI -0.9%, -0.8%). No changes were observed during the restriction or recovery periods.

For parenteral opioids, the median prevalence was 0.4 per 1000 per month pre-COVID-19. However, there were large increases in prescribing in March-May 2020, including a 18.0% (95%CI 6.1%, 31.2%) increase in March, a 89.4% (95%CI 76.0%, 103.8%) increase in April, and a 16.8% (95%CI 8.3%, 26.0%) increase in May. Even after accounting for these temporary increases, weak evidence of a positive level shift was observed during lockdown (10.7%, 95%CI -0.4%, 23.1%), which reduced during the recovery period (-8.4%, 95%CI -14.6%, -1.8%).

### Changes in opioid prescribing by subgroup

#### People living in care homes

There was a median of 155,943 registered adult patients living in a care home per month. Prior to the start of COVID-19 period, a median of 182.4 people were prescribed an opioid per 1000 patients in a care home, which declined by 0.2% per month (95%CI -0.3%, -0.2%) (**Figure 2**, **Table 2**). An increase in prevalent prescribing was observed in March (2.9%, 95%CI 0.3%, 5.6%) and April (13.3%, 95%CI 11.2%, 15.4%), and there was a small negative level shift during the recovery period (-1.5%, 95%CI -3.0%, -0.01%). Prescribing of high dose, long-acting opioids also declined by 1.3% per month (95%CI -1.4%, -1.1%), with an increasing slope starting in the recovery period (1.5%, 95%CI 1.3%, 1.8%).

**Figure 2.**
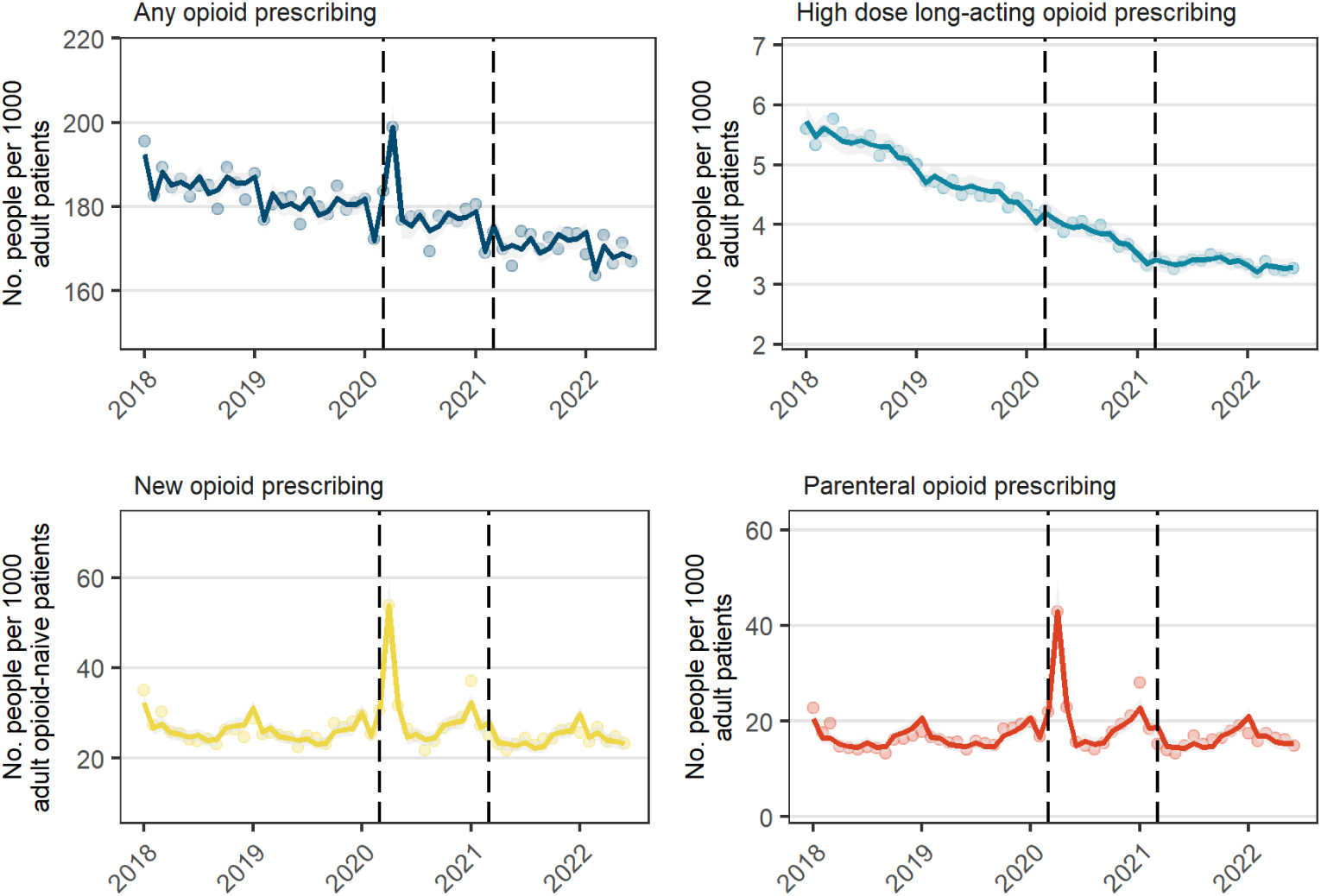
Number of people prescribed opioids per month (Jan 2018 to Mar 2022) among registered patients living in care homes. Solid lines are fitted values, dots are observed values, and vertical dashed lines represent start of restriction period (Mar 2020) and recovery period (Apr 2021).

Median new opioid prescribing was 25.4 per 1000 opioid-naïve patients in a care home pre-COVID-19 and was stable. Increases in new prescribing were observed in April (112.5%, 95%CI 92.2%, 134.9%) and May (26.0%, 95%CI 14.6%, 38.5%). After accounting for these changes, no other changes were observed during the lockdown period. There was a -10.2% level shift (95%CI -18.7%, -0.7%) in new prescribing starting in the recovery period.

Prescribing of parenteral opioids was much higher in care homes than in the general population (16.2 per 1000 pre-COVID-19) and there were large increases in prescribing in Mar-May 2020, including a 35.9% (95%CI 11.6%, 65.5%) increase in March, 186.3% (95%CI 153.1%, 223.9%) in April, and 54.2% (95%CI 37.0%, 73.7%) in May. Aside from these temporary changes, there was also weak evidence of a level shift during the recovery period of -14.3% (95%CI -26.7%, 0.2%).

#### Demographic subgroups

Demographic variation in prevalent and new opioid prescribing by month (**Figure 3**, **Figure 4**) mirrored those observed in **Table 1**. During the restriction period, there was a negative shift in overall opioid prescribing for people aged 18-29 years (-5.2%, 95%CI -8.9%, -1.4%), but no change for all other age groups (**Figure 5**, **Supplementary Table 2**). Other differences include a decrease in people with Asian or Asian British ethnicity (-5.7%, 95%CI -8.5%, -2.8%), Other ethnicity (-4.2%, 95%CI -7.2%, -1.2%) and people living in London (-5.8%, 95%CI -8.9%, -2.5%). These decreases did not reverse during the recovery period. For people aged 18-29 years, there was a further level shift of -4.2% (95%CI -7.2%, -1.2%) during the recovery period. There was little variation by sex or IMD decile.

**Figure 3.**
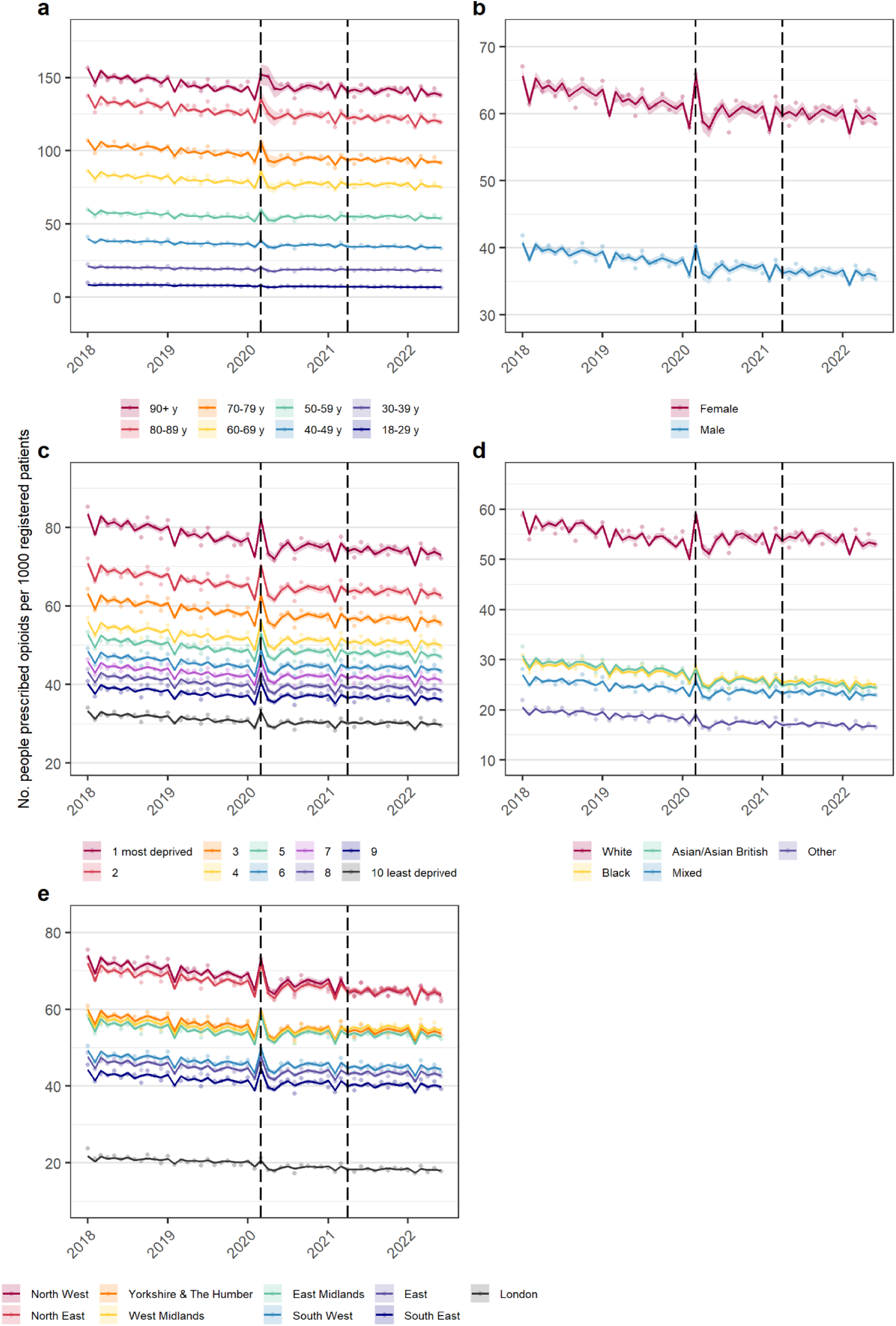
Number of people prescribed opioids per month (Jan 2018 to Mar 2022) among all registered adult patients, by: a. age category; b. sex; c. IMD decile; d. ethnicity; e. region. Solid lines are fitted values, dots are observed values, and vertical black lines represent the start of restriction period (Mar 2020) and recovery period (Apr 2021).

**Figure 4.**
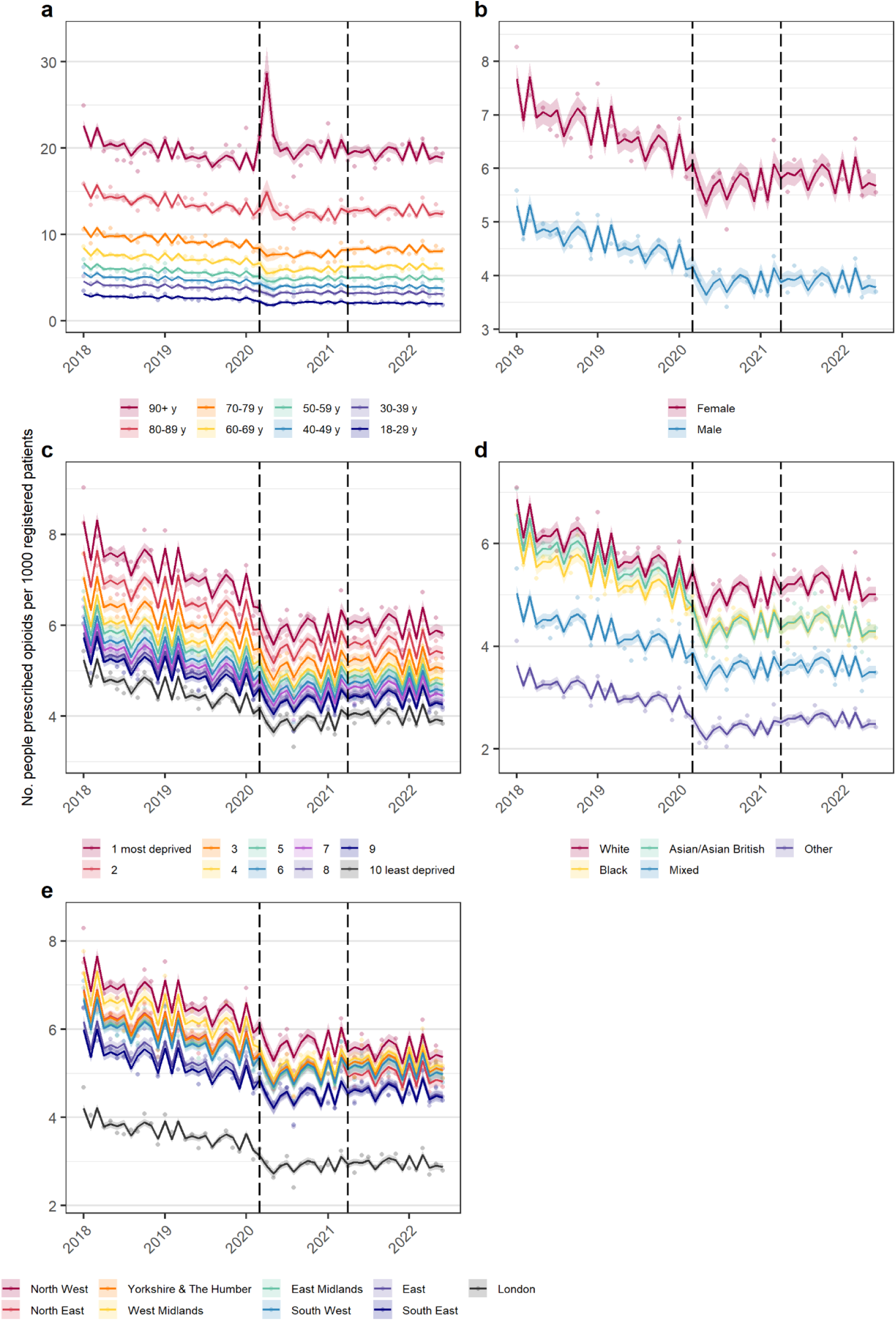
Number of people newly prescribed opioids per month (Jan 2018 to Mar 2022), among all registered adult patients, by: a. age category; b. sex; c. IMD decile; d. ethnicity; e. region. Solid lines are fitted values, dots are observed values, and vertical black lines represent the start of restriction period (Mar 2020) and recovery period (Apr 2021).

**Figure 5.**
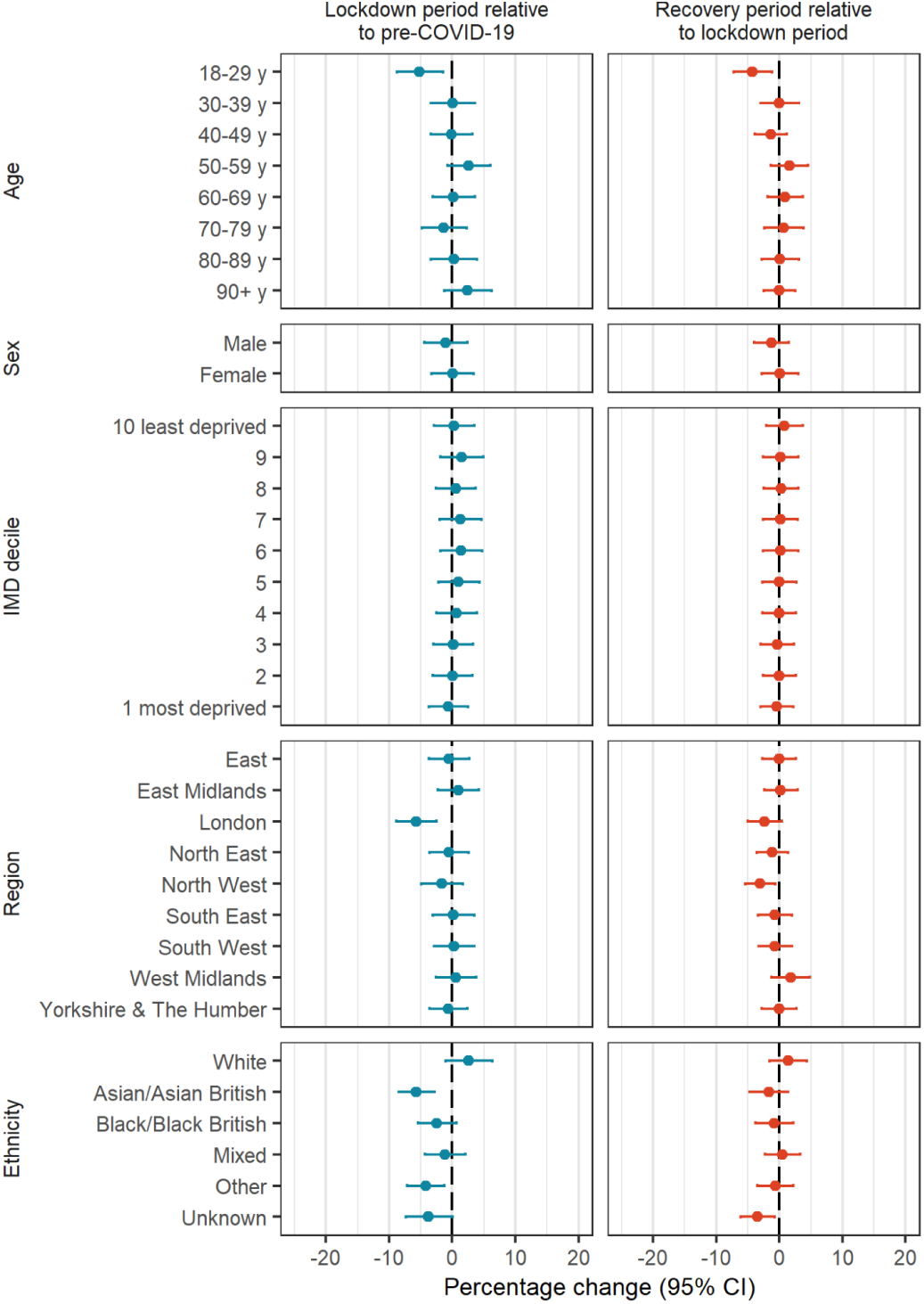
Changes in number of people prescribed opioids during the restriction and recovery periods by demographics, adjusted for long-term trends and seasonality

For new opioid prescribing, no significant differences were observed by sex. The change associated with the restriction period varied most dramatically by age (**Figure 6**, **Supplementary Table 3**). There were negative shifts in new prescribing ranging from -7.0% to -13.0% in age categories <80 years; these decreases reversed in the recovery period for people aged 60-79 years, but not in younger age groups. A dramatic increase in new prescribing was observed in April 2020 for people 90+ years, which was not observed for other age groups. For most other demographic categories, there were similar decreases in new prescribing during the restriction period, with minimal evidence of rebounding during the recovery period.

**Figure 6.**
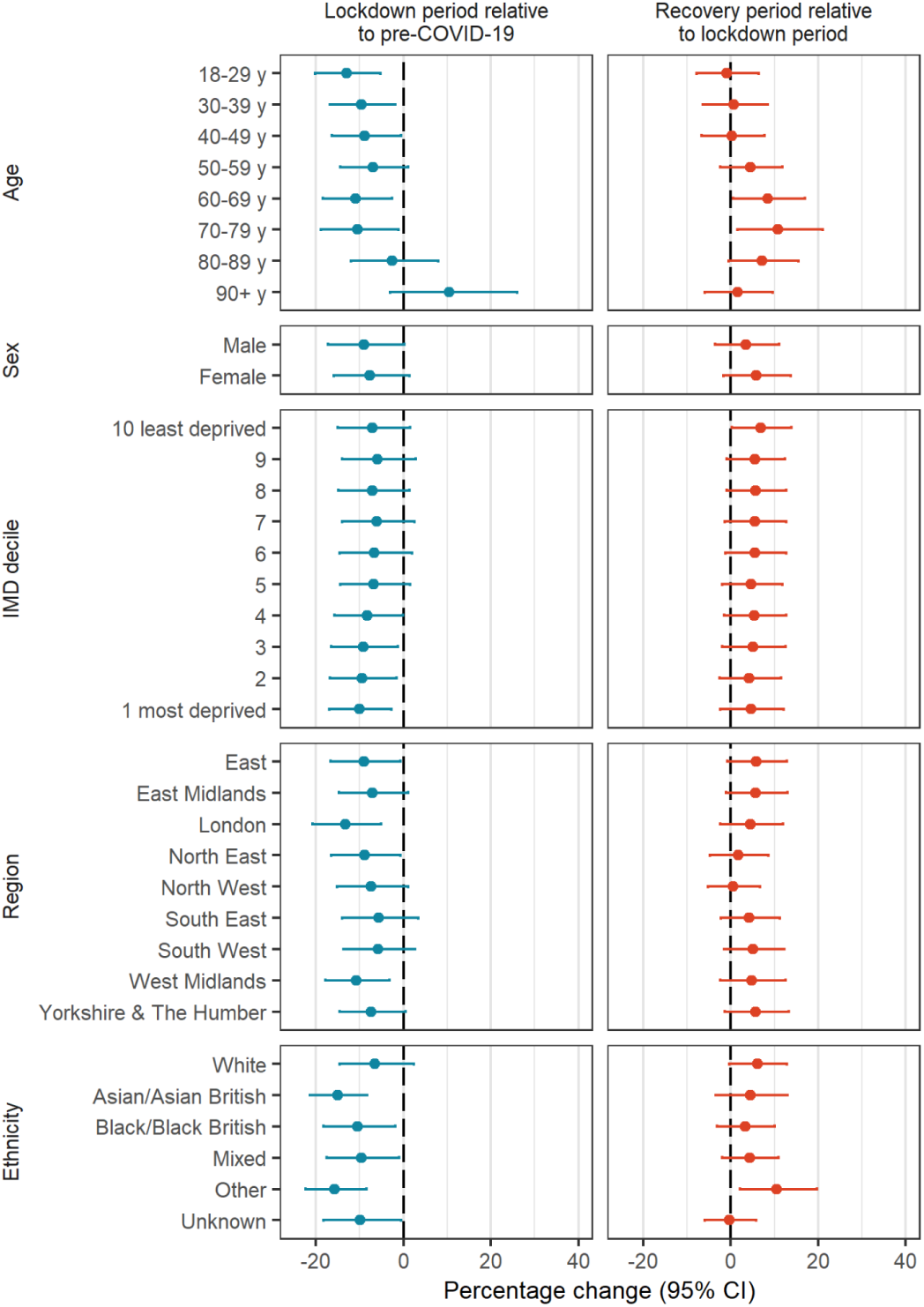
Changes in number of people newly opioid prescribing during the restriction and recovery periods by demographics, adjusted for long-term trends and seasonality

## Discussion

### Summary

While the COVID-19 pandemic was associated with minimal changes in prevalent opioid prescribing in England, we found sustained decreases in people newly prescribed opioids, with weak evidence that this may have reversed slightly during the recovery period. And while our findings confirm previously identified differences in opioid prescribing by socioeconomic deprivation, ethnicity, and geography,(7,8) we found only minimal differences in how the pandemic impacted on opioid prescribing by sex, IMD decile, region, and ethnicity. The exception was older people and people living in care homes where the prescribing of parenteral opioids coincided with increases in COVID-19 morbidity and mortality, strongly suggesting use to treat end-of-life COVID-19 symptoms such as breathlessness.(19)

### Findings in context

Our work confirms the continuation of the downward trend in opioid prescribing in England starting in 2016 observed in aggregate prescribing data(8), coinciding with a policy focus on reducing opioid prescribing for chronic non-cancer pain.(5,20) However, few studies have evaluated changes to the number of people prescribed opioids during the pandemic, with most focusing on number of prescriptions or dispensings. Consistent with our findings, one study found a decrease in new opioid users among people with certain musculoskeletal or rheumatic conditions, but not in the number of overall prescriptions.(13) This reduction was attributed by those authors to caution from GPs in newly prescribing opioids during the pandemic when monitoring was more difficult. Early in the pandemic, there was also a decrease in elective medical procedures that often require prescribed analgesia(21); however, opioids would typically be supplied by the hospital and are not captured in our data. Therefore, reduced new opioid prescribing is more likely related to decreased primary care contacts and fewer opportunities for opioids to be initiated.(22).

People in care homes were greatly impacted by the pandemic and in the first 12 weeks one third of all deaths in care homes were attributable to COVID-19.(23) The observed spikes in parenteral opioid prescribing coincide with the peak in mortality (both COVID-19 and non-COVID-19) (**Supplementary Figure 2**)(24) strongly suggesting this is related to treatment of people at the end of life. Another OpenSAFELY study on end-of-life care found that 21% who died of COVID-19 received medication to manage symptoms.(25) A similar pattern was observed with antipsychotic prescribing to people in care homes(26).

While opioid prescribing for treatment of patients at the end-of-life is best practice,(27) other studies have identified increases in prescribing of sedating medicines (opioids, antidepressants, antipsychotics, benzodiazepines) to people in care homes,(9,26) to deal with the psychological symptoms resulting from increased social isolation during the pandemic.(28) We observed high monthly rates of opioid prescribing (18%) to people in care homes even prior to the COVID-19 pandemic, and this population is especially at risk of harms such as falls and respiratory depression.

### Strengths and limitations

Our data capture primary care records for approximately 40% of patients registered with a practice in England via the OpenSAFELY platform, and these data are representative of the wider English population.(29) This study is the largest to quantify person-level changes in opioid prescribing during the COVID-19 pandemic in England, allowing us to identify changes by demographic groups and in new prescribing patterns, which is not possible with aggregate data.

While it is important to understand how opioid prescribing has changed, it is limited in what it tells us about the quality of prescribing as we don’t know the reason for prescribing, as is common with all large database EHR studies. Furthermore, we did not have information on prescribed dose or duration, which could help identify more nuanced changes such as dose escalation. Our data only includes prescriptions carried out in primary care not in secondary care (e.g., hospitals), although we have advocated strongly for improvements in NHS hospital data infrastructure to make this possible.(30) Lastly, there are no definitive data on people living in care homes in England, and we have relied on a previously established algorithm(17) which produces similar estimates to ONS data.(31)

### Policy implications

A 2019 report by Public Health England raised concerns about the high prevalence of opioid prescribing, and emphasised the need for quality and contemporary and detailed data on predictors of long term use and dependence.(1) NHS England, the body with national responsibility for care, issued instructions to improve opioid use in 2023, highlighting the need for better use of data to prevent and reduce opioid harm.(32) This study demonstrates how OpenSAFELY can be used for fine grain analysis of opioid prescribing in line with key recommendations. We are developing tools to facilitate near real-time audit and feedback in the context of rapidly evolving pressures on the health service readily extendable to other clinical and challenges and can include any measures on opioids needed to support NHS England’s ambition on safe opioid use.

### Future research

In addition to providing near real time feedback for the health system on opioid use, OpenSAFELY could support a range of other analyses. The COVID-19 pandemic led to worsening of the backlog in elective procedures with nearly 7.0 million people on waiting lists as of August 2022 (31) putting people at increased risk of long-term opioid use; quantifying this impact in a large, representative sample is needed. We are actively incorporating waiting list data into OpenSAFELY to support this work. Continued vigilance of opioid use in people experiencing long-term symptoms of COVID-19, which includes joint and musculoskeletal pain,(33) is also warranted.

## Conclusion

We found little change in overall opioid prescribing apart from temporary changes at the start of the first lockdown, with reductions in new opioid prescribing which were sustained until the end of the study period. Conversely, we observed a substantial increase in parenteral opioid prescribing and new opioid prescribing for people living in care homes, coinciding with the peak in COVID-19 morbidity and mortality and likely representing use to treat end-of-life COVID-19 symptoms. Continued monitoring of changes in opioid prescribing in light of the ongoing pandemic and potential long-term COVID-19 symptoms is warranted.

## Data Availability

https://github.com/opensafely/opioids-covid-research

## Administrative

## Acknowledgements

We are very grateful for all the support received from the TPP Technical Operations team throughout this work, and for generous assistance from the information governance and database teams at NHS England and the NHS England Transformation Directorate.

Members of the The OpenSAFELY Collaborative include: Sebastian CJ Bacon, Lucy Bridges, Benjamin FC Butler-Cole, Simon Davy, Iain Dillingham, David Evans, Louis Fisher, Amelia Green, Ben Goldacre, Liam Hart, George Hickman, Peter Inglesby, Steven Maude, Jessica Morley, Amir Mehrkar, Thomas O’Dwyer, Rebecca M Smith, Pete Stokes, Tom Ward, Jon Massey, Milan Wiedemann, Christopher Bates, Jonathan Cockburn, Sam Harper, Frank Hester, John Parry.

## Author contributions

Conceptualization: ALS, BMK, AJW; Data curation: ALS, CA, SCJB, CB, BG, JM, PI; Formal analysis: ALS, CA, MW; Funding acquisition: BG; Investigation: ALS; Methodology: ALS, BMK, CW, RC; Project administration: AJW, BMK, AM, BG; Resources: AJW, RW, SCJB, BG; Software: ALS, CA, MW, JM, PI; Supervision: AJW, BMK, AM, BG; Validation: ALS, CA, MW; Visualisation: ALS; Writing - original draft: ALS; Writing - review & editing: All authors. ALS, JM, PI directly accessed and verified the underlying data reported in the manuscript.All authors gave final approval of the version to be published and agree to be accountable for all aspects of the work. All authors confirm that they had full access to all the data in the study and accept responsibility to submit for publication.

## Transparency

The lead author affirms that the manuscript is an honest, accurate, and transparent account of the study being reported; that no important aspects of the study have been omitted; and that any discrepancies from the study as originally planned have been explained.

## Information governance

NHS England is the data controller of the NHS England OpenSAFELY COVID-19 Service; TPP is the data processor; all study authors using OpenSAFELY have the approval of NHS England.(1) This implementation of OpenSAFELY is hosted within the TPP environment which is accredited to the ISO 27001 information security standard and is NHS IG Toolkit compliant.(2)

Patient data has been pseudonymised for analysis and linkage using industry standard cryptographic hashing techniques; all pseudonymised datasets transmitted for linkage onto OpenSAFELY are encrypted; access to the NHS England OpenSAFELY COVID-19 service is via a virtual private network (VPN) connection; the researchers hold contracts with NHS England and only access the platform to initiate database queries and statistical models; all database activity is logged; only aggregate statistical outputs leave the platform environment following best practice for anonymisation of results such as statistical disclosure control for low cell counts.(3)

The service adheres to the obligations of the UK General Data Protection Regulation (UK GDPR) and the Data Protection Act 2018. The service previously operated under notices initially issued in February 2020 by the Secretary of State under Regulation 3(4) of the Health Service (Control of Patient Information) Regulations 2002 (COPI Regulations), which required organisations to process confidential patient information for COVID-19 purposes; this set aside the requirement for patient consent.(4) As of 1 July 2023, the Secretary of State has requested that NHS England continue to operate the Service under the COVID-19 Directions 2020.(5) In some cases of data sharing, the common law duty of confidence is met using, for example, patient consent or support from the Health Research Authority Confidentiality Advisory Group.(6)

Taken together, these provide the legal bases to link patient datasets using the service. GP practices, which provide access to the primary care data, are required to share relevant health information to support the public health response to the pandemic, and have been informed of how the service operates.

## Supplementary information

**Supplementary Figure 1.**
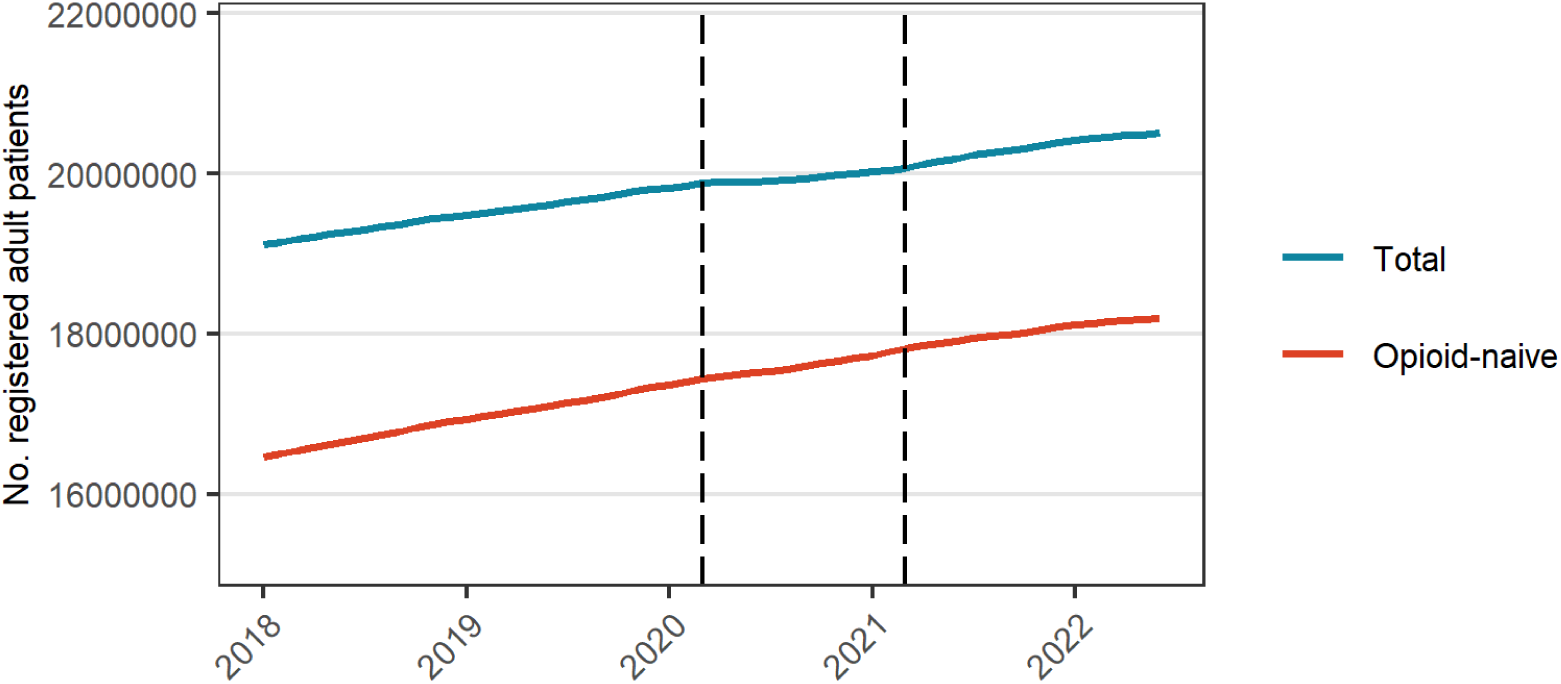
Number of registered adult patients by month during the study period. Opioid-naive is defined as people with an opioid prescription in the past year. Vertical dash lines represent the start of the restriction (Mar 2020) and recovery (Apr 2021) periods.

**Supplementary Figure 2.**
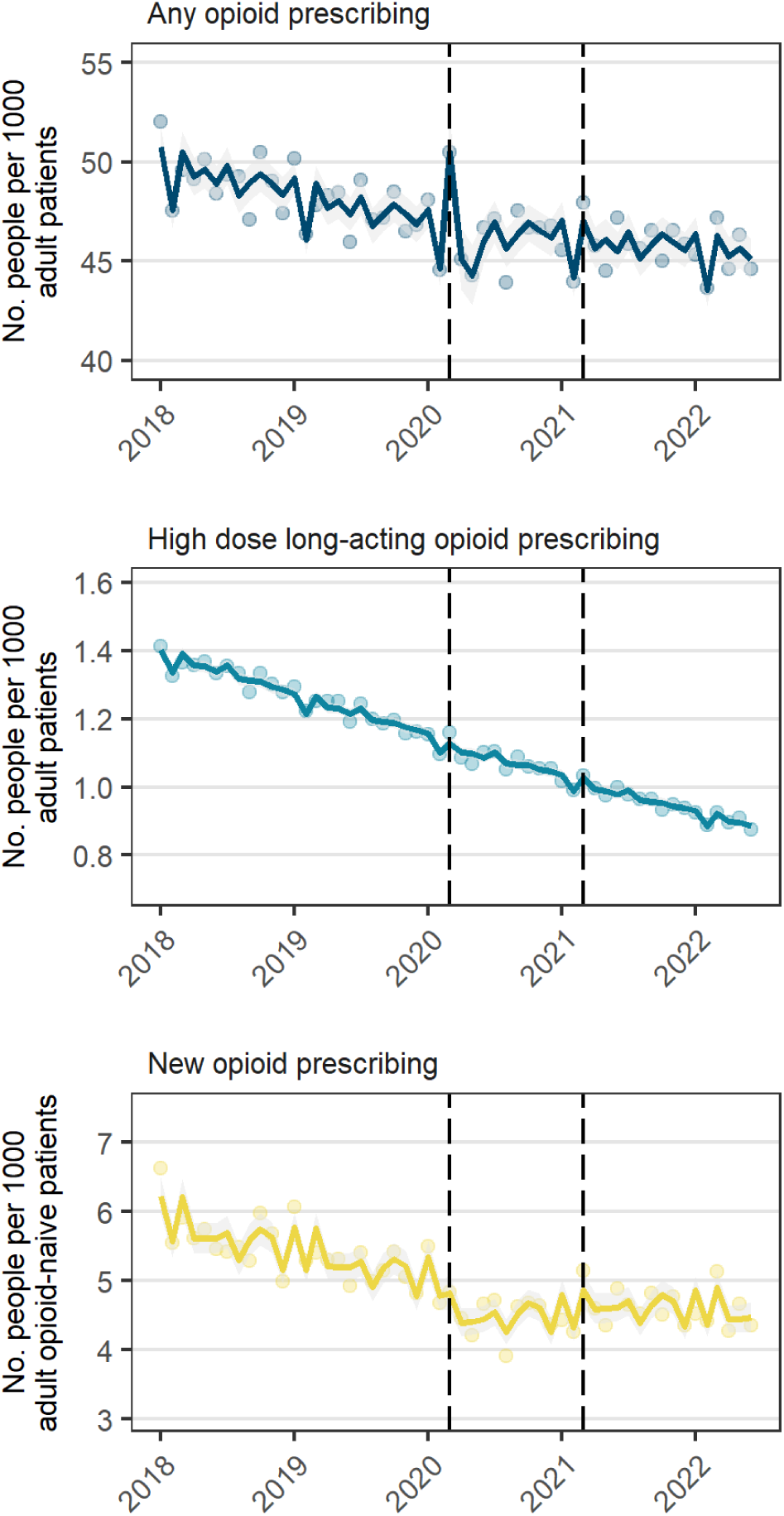
Number of people prescribed opioids per month (Jan 2018 to June 2022) among all registered adult patients without a history of cancer. Solid lines are fitted values, dots are observed values, and vertical dashed lines represent start of restriction period (Mar 2020) and recovery period (Apr 2021).

**Supplementary Table 1.** Relative changes in number of people prescribed opioids per 1000 population during the restriction (Mar 2020-Mar 2021) and recovery (Apr 2021-Jun 2022) periods among all registered adult patients without a history of cancer.

|  | Pre-COVID-19<br>monthly slope<br>(%, 95% CI) | Changes during restriction period relative to pre-COVID-19 |  |  |  |  | Changes during recovery period<br>relative to restriction period |  |
| --- | --- | --- | --- | --- | --- | --- | --- | --- |
|  |  | Level shift<br>(%, 95% CI) | Change in slope<br>(%, 95% CI) | March 2020 (%,<br>95% CI) | April 2020 (%,<br>95% CI) | May 2020 (%,<br>95% CI) | Level shift (%,<br>95% CI) | Change in slope<br>(%, 95% CI) |
| **Full adult population** |  |  |  |  |  |  |  |  |
| Any opioid | -0.3 (-0.3, -0.2) | -0.6 (-3.5, 2.3) | 0.2 (-0.1, 0.6) | 7.0 (3.3, 10.9) | -2.2 (-4.4, 0.07) | -4.9 (-7.9, -1.8) | -0.6 (-3.5, 2.3) | -0.1 (-0.4, 0.3) |
| New opioid | -0.6 (-0.8, -0.5) | -10.2 (-13.3,<br>-7.1) | 0.7 (0.3, 1.1) | * | * | * | 4.1 (-0.8, 9.3) | -0.4 (-0.8, 0.1) |
| High dose long-acting<br>opioid | -0.8 (-0.8, -0.8) | -1.7 (-3.3, 0.02) | 0.03 (-0.1, 0.2) | * | * | * | -1.1 (-2.4, 0.2) | -0.05 (-0.2, 0.1) |

\*Not included in model

**Supplementary Table 1.**
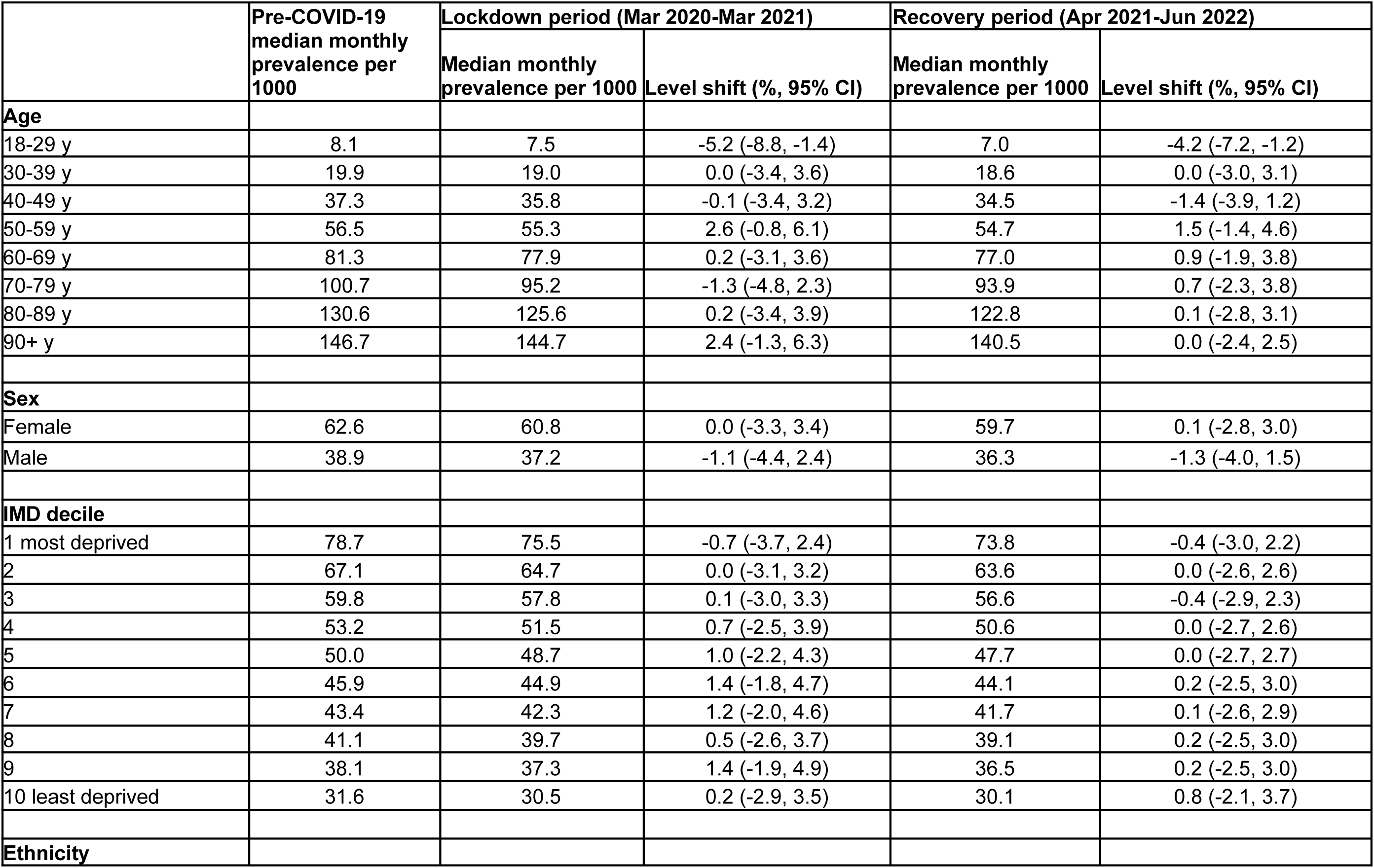
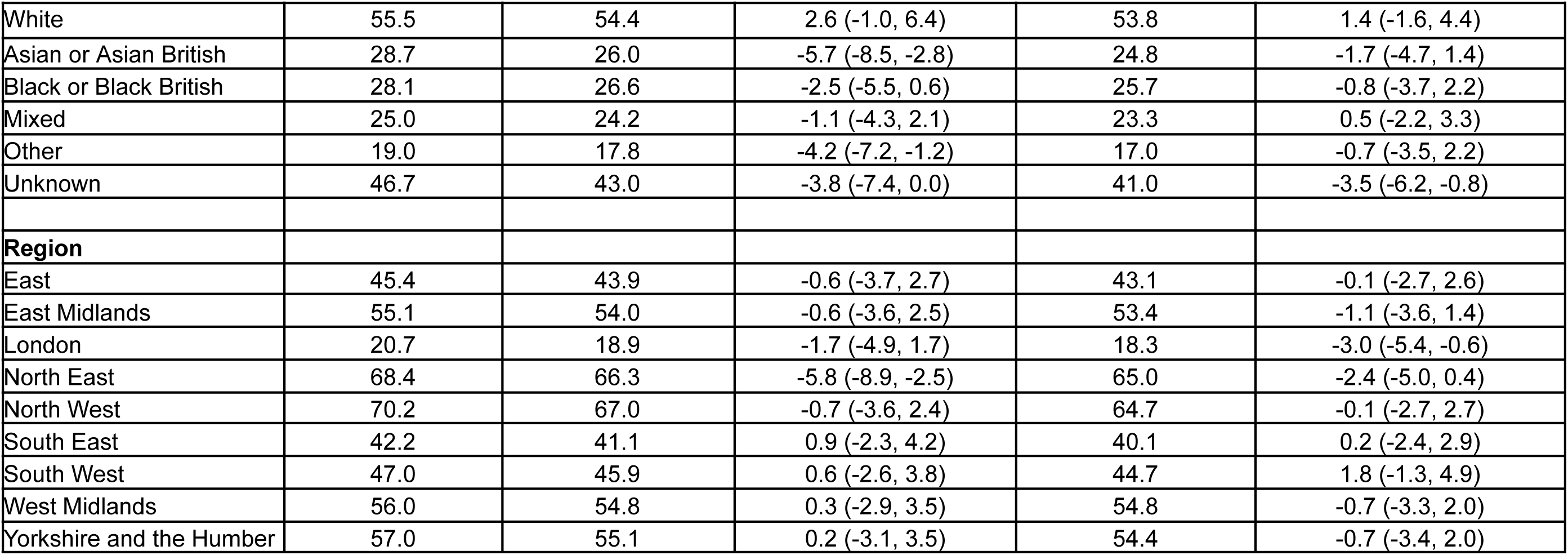
Changes in number of people prescribed opioids changes during lockdown and recovery periods by demographic categories.

**Supplementary Table 2.** Changes in number of people newly prescribed opioids during lockdown and recovery periods by demographic categories.

|  | Pre-COVID-19<br>median monthly<br>prevalence per 1000 | Lockdown period (Mar 2020-Mar 2021) |  | Recovery period (Apr 2021-Jun 2022) |  |
| --- | --- | --- | --- | --- | --- |
|  |  | Median monthly<br>prevalence per<br>1000 | Level shift (95% CI) | Median monthly<br>prevalence per<br>1000 | Level shift (95% CI) |
| Age |  |  |  |  |  |
| 18-29 y | 2.7 | 2.2 | -13.0 (-20.2, -5.2) | 2.1 | -1.0 (-7.8, 6.4) |
| 30-39 y | 4.0 | 3.4 | -9.6 (-16.8, -1.8) | 3.1 | 0.7 (-6.4, 8.4) |
| 40-49 y | 4.8 | 4.1 | -8.8 (-16.4, -0.5) | 3.9 | 0.3 (-6.6, 7.7) |
| 50-59 y | 5.7 | 4.9 | -7.0 (-14.5, 1.1) | 4.9 | 4.4 (-2.4, 11.8) |
| 60-69 y | 7.4 | 5.9 | -10.9 (-18.5, -2.6) | 6.2 | 8.4 (0.6, 16.9) |
| 70-79 y | 9.5 | 7.7 | -10.4 (-18.9, -1.1) | 8.2 | 10.8 (1.6, 21.0) |
| 80-89 y | 13.7 | 12.3 | -2.6 (-12.0, 7.9) | 12.5 | 7.2 (-0.5, 15.4) |
| 90+ y | 19.5 | 20.2 | 10.5 (-3.0, 25.9) | 19.4 | 1.5 (-5.9, 9.6) |
| Sex |  |  |  |  |  |
| Male | 6.7 | 5.8 | -7.7 (-15.9, 1.4) | 5.8 | 5.7 (-1.6, 13.7) |
| Female | 4.7 | 3.9 | -9.0 (-17.3, 0.2) | 3.9 | 3.5 (-3.6, 11.1) |
| IMD decile |  |  |  |  |  |
| 1 most deprived | 7.2 | 6.1 | -10.1 (-17.0, -2.7) | 5.9 | 4.6 (-2.4, 12.1) |
| 2 | 6.7 | 5.7 | -9.5 (-16.8, -1.6) | 5.5 | 4.2 (-2.6, 11.5) |
| 3 | 6.2 | 5.3 | -9.2 (-16.5, -1.2) | 5.1 | 5.0 (-2.0, 12.5) |
| 4 | 5.8 | 4.9 | -8.2 (-15.7, -0.1) | 4.9 | 5.4 (-1.5, 12.7) |
| 5 | 5.7 | 4.8 | -6.9 (-14.5, 1.4) | 4.8 | 4.7 (-2.0, 11.9) |
| 6 | 5.4 | 4.7 | -6.7 (-14.7, 1.9) | 4.7 | 5.5 (-1.2, 12.7) |
| 7 | 5.3 | 4.6 | -6.1 (-14.1, 2.6) | 4.6 | 5.4 (-1.3, 12.7) |
| 8 | 5.1 | 4.3 | -7.1 (-14.9, 1.4) | 4.4 | 5.6 (-0.9, 12.6) |
| 9 | 5.0 | 4.3 | -6.0 (-14.1, 2.9) | 4.3 | 5.5 (-0.9, 12.4) |
| 10 least deprived | 4.6 | 3.9 | -7.2 (-15.1, 1.5) | 3.9 | 6.8 (0.2, 13.9) |

| **Ethnicity** |  |  |  |  |  |
| --- | --- | --- | --- | --- | --- |
| White | 5.9 | 5.1 | -6.5 (-14.6, 2.4) | 5.1 | 6 (-0.4, 12.9) |
| Asian or Asian British | 5.7 | 4.5 | -15.1 (-21.4, -8.3) | 4.4 | 4.4 (-3.4, 12.9) |
| Black or Black British | 5.4 | 4.5 | -10.5 (-18.3, -1.8) | 4.5 | 3.2 (-3.2, 10.1) |
| Mixed | 4.3 | 3.6 | -9.6 (-17.5, -1.0) | 3.6 | 4.3 (-1.9, 10.9) |
| Other | 3.1 | 2.4 | -15.7 (-22.3, -8.5) | 2.5 | 10.5 (2.1, 19.7) |
| Unknown | 5.1 | 4.3 | -9.9 (-18.3, -0.6) | 4.0 | -0.3 (-6.0, 5.8) |
| **Region** |  |  |  |  |  |
| East | 5.4 | 4.5 | -9.0 (-16.7, -0.7) | 4.6 | 5.8 (-0.8, 12.9) |
| East Midlands | 5.9 | 5.1 | -8.9 (-16.5, -0.6) | 5.1 | 1.6 (-4.8, 8.5) |
| London | 3.7 | 2.9 | -7.4 (-15.2, 1.1) | 2.9 | 0.5 (-5.3, 6.7) |
| North East | 6.0 | 5.1 | -13.3 (-20.7, -5.1) | 5.0 | 4.5 (-2.3, 11.9) |
| North West | 6.7 | 5.7 | -7.4 (-14.6, 0.5) | 5.1 | 5.7 (-1.4, 13.2) |
| South East | 5.2 | 4.5 | -7.2 (-14.8, 1.1) | 4.5 | 5.7 (-1.1, 12.9) |
| South West | 5.8 | 5.0 | -10.8 (-17.8, -3.2) | 5.1 | 4.8 (-2.4, 12.6) |
| West Midlands | 6.4 | 5.3 | -5.8 (-13.7, 2.7) | 5.3 | 5.1 (-1.5, 12.2) |
| Yorkshire and the Humber | 6.0 | 5.2 | -5.7 (-14.0, 3.5) | 5.2 | 4.2 (-2.2, 11.1) |

**Supplementary Figure 2.**
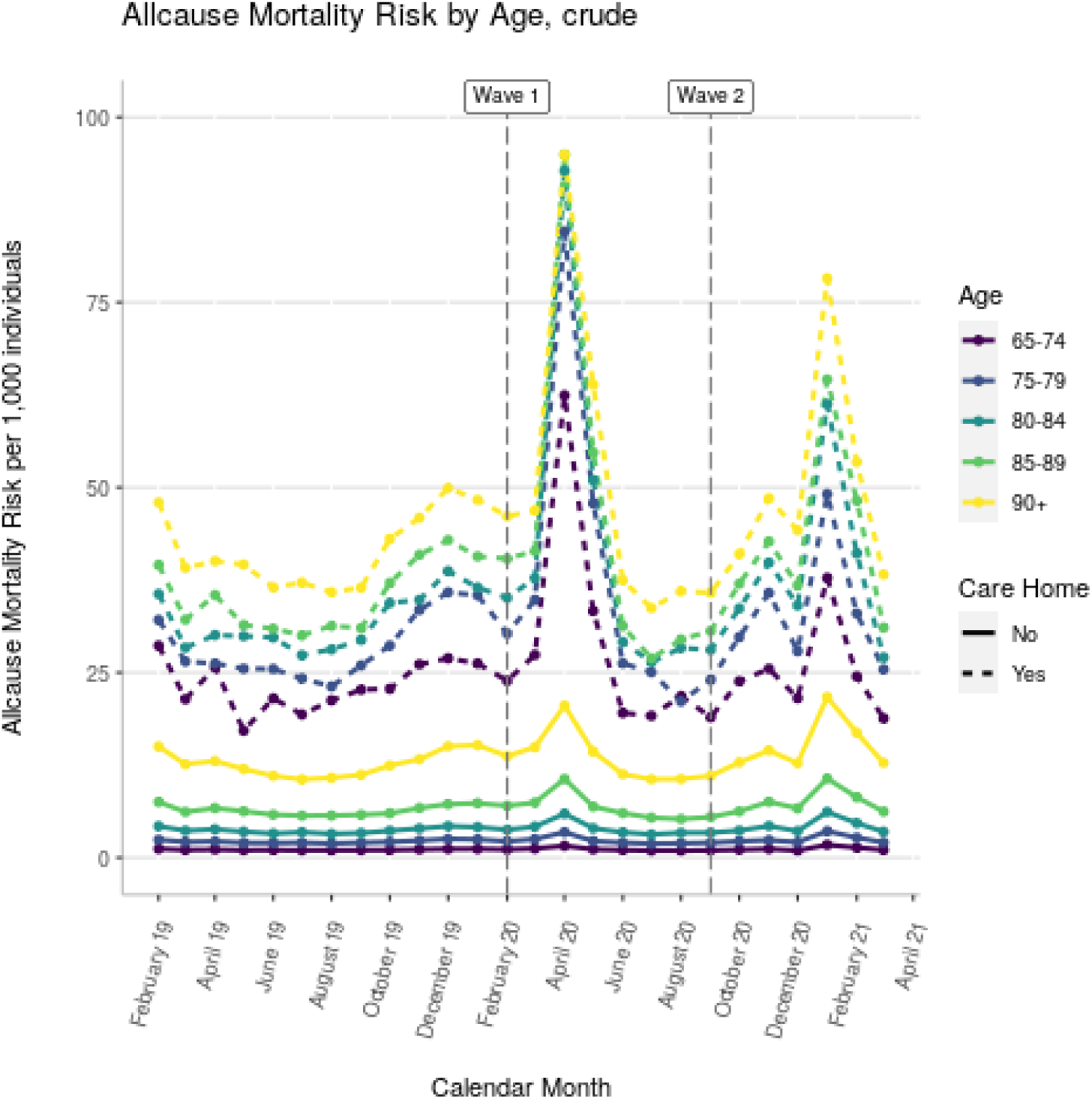
All cause mortality by age group and care home status, from Schultze et al. Mortality among Care Home Residents in England during the first and second waves of the COVID-19 pandemic: an observational study of 4.3 million adults over the age of 65. *Lancet Reg Health Eur*. 2022 Mar; 14: 100295. doi: 10.1016/j.lanepe.2021.100295

